# Testing the Ability of Convolutional Neural Networks to Learn Radiomic Features

**DOI:** 10.1101/2020.09.19.20198077

**Authors:** Ivan S. Klyuzhin, Yixi Xu, Anthony Ortiz, Juan Lavista Ferres, Ghassan Hamarneh, Arman Rahmim

**Affiliations:** Department of Integrative Oncology, BC Cancer Research Institute, Vancouver, BC, Canada; Department of Radiology, University of British Columbia, Vancouver, BC, Canada; AI for Health, Microsoft, Redmond, WA, USA; Department of Computing Science, Simon Fraser University, Burnaby, BC, Canada; Department of Physics and Astronomy, University of British Columbia, Vancouver, BC, Canada

**Keywords:** Deep learning, radiomics, cancer, medical imaging, image analysis

## Abstract

**Background and Objective:** Radiomics and deep learning have emerged as two distinct approaches to medical image analysis. However, their relative expressive power remains largely unknown. Theoretically, hand-crafted radiomic features represent a mere subset of features that neural networks can approximate, thus making deep learning a more powerful approach. On the other hand, automated learning of hand-crafted features may require a prohibitively large number of training samples. Here we directly test the ability of convolutional neural networks (CNNs) to learn and predict the intensity, shape, and texture properties of tumors as defined by standardized radiomic features.

**Methods:** Conventional 2D and 3D CNN architectures with an increasing number of convolutional layers were trained to predict the values of 16 standardized radiomic features from real and synthetic PET images of tumors, and tested. In addition, several ImageNet-pretrained advanced networks were tested. A total of 4000 images were used for training, 500 for validation, and 500 for testing.

**Results:** Features quantifying size and intensity were predicted with high accuracy, while shape irregularity and heterogeneity features had very high prediction errors and generalized poorly. For example, mean normalized prediction error of tumor diameter with a 5-layer CNN was 4.23 ± 0.25, while the error for tumor sphericity was 15.64 ± 0.93. We additionally found that learning shape features required an order of magnitude more samples compared to intensity and size features.

**Conclusions:** Our findings imply that CNNs trained to perform various image-based clinical tasks may generally under-utilize the shape and texture information that is more easily captured by radiomics. We speculate that to improve the CNN performance, shape and texture features can be computed explicitly and added as auxiliary variables to the networks, or supplied as synthetic inputs.

## INTRODUCTION

Quantitative pattern analysis in radiological images can be used to assess tumor phenotype as well as micro- and macro-environmental conditions *(1)*. For example, larger and more heterogeneous tumors as measured from positron emission tomography (PET) and computed tomography (CT) images have been found to be generally more aggressive and more resilient to treatment *(2, 3, 4)*, while more irregular tumor shapes have been associated with a lower probability of complete response *(5)*. In addition, tumor-specific texture characteristics can be used for automated lesion detection and segmentation *(6, 7)*. Given these findings, there have been considerable efforts to develop novel pattern analysis methods for medical imaging of cancer and other diseases *(8, 9, 10, 11)*.

Two distinct approaches have emerged: radiomics and deep learning. Radiomics-based methods utilize hand-crafted features that are intended to capture various properties of the tumor, e.g. its shape and texture *(1, 12, 13)*. Various radiomic features have been found to be significant predictors of disease-free survival and response to therapy *(14-17)*. Deep learning methods in medical imaging typically utilize convolutional neural networks (CNN) trained in an end-to-end fashion, with images serving as inputs and clinical metrics as targets. In the process of training, relevant low- and high-level image features become automatically and implicitly encoded in the layers of the network *(18)*. Thus, deep learning methods eliminate the need for feature design and selection, and can potentially forego the need for image segmentation *(16)*.

Recent reports of human-level cancer detection performance by CNNs *(19-22)* may suggest that emphasis in method development should be placed on deep learning, rather than radiomics. According to the universal approximation theorem *(23, 24)*, hand-crafted radiomic features represent a subset of functions that CNNs can approximate, seemingly obviating the practice of using explicit radiomics for predictive tasks. The problem, however, is that the theorem does not provide any bounds on the required number of neurons to approximate a function: the necessary number of CNN layers or nodes to match the power of a hand-crafted feature may well be impractical. Sample complexity is another concern: the number of samples required to learn a particular feature may be unrealistic, or vary substantially between the features, leading to significant biases in learning of different kinds of information (e.g. texture versus shape) *(25)*. Thus, in some scenarios, it may be more efficient and effective to use radiomic features instead of neural networks.

In the present work, we directly test the ability of CNNs to learn hand-crafted and standardized radiomic features, and measure the sample complexity for different features. To that end, we train simple CNN architectures with a progressively larger number of convolutional layers (up to nine), and several advanced ImageNet-pretrained architectures, to predict the explicitly-computed values of radiomic features. A poor prediction accuracy for a particular feature would imply that common CNN architectures may be unable to effectively capture and use the corresponding type of information (for a given number of samples and network size). Training and testing are done using 2 sets of real 2D PET images comprised of lymphoma and head and neck cancer lesions, as well as 2 sets of synthetic 2D and 3D lesion images.

## MATERIALS AND METHODS

### Acquired Images

Images of real tumors were extracted from two datasets. The first dataset was obtained locally and contained whole-body ^18^F-fludeoxyglucose (FDG) PET/CT volumes of patients with primary mediastinal B-cell lymphoma who were treated with R(rituximab)-CHOP. Collection of human imaging data was approved by the University of British Columbia - BC Cancer Research Ethics Board (UBC BC Cancer REB), and all subjects gave informed written consent. A total of 126 volumes from 69 unique subjects were available for the study, acquired on a GE Discovery 690 scanner at baseline, mid-, and post-treatment (after 3-6 chemotherapy cycles). The injected activity ranged from ∼280 to ∼450 MBq and the PET scans were performed 60 minutes after the injection. The images were reconstructed iteratively with point spread function modeling, but without time-of-flight modeling (GE “VPHDS” reconstruction). The axial dimensions of reconstructed images were 192×192, with isotropic voxel size (3.64 mm)^3^. The voxel intensities were normalized to represent the standardized uptake values (SUVs). In all images, primary tumors were manually delineated in 3D by a nuclear medicine physician.

The second dataset was obtained from the publicly-available HECKTOR challenge (head and neck tumor segmentation and outcome prediction), which includes 224 FDG PET/CT images of patients with head and neck cancer acquired at multiple centers (https://www.aicrowd.com/challenges/miccai-2021-hecktor). The images from this dataset were re-sampled to have the same voxel size as the lymphoma dataset.

The number of volume images in both datasets was insufficient to robustly train 3-dimensional CNNs. Thus, we generated corresponding much larger sets of real 2D images (and masks) by slicing the tumor volumes with regularly-spaced axial, sagittal, and coronal planes. The slicing planes were 2 voxels apart in each dimension. The resulting full-size slices were cropped to the size 48×48 pixels (without interpolation), such that the lesion centroid was located in the center of the cropped image. After excluding images where the lesion area was less than 50 pixels and the maximum SUV was less than 3.0, this yielded 2008 2D lesion images of lymphoma and 1432 images of head and neck cancer. Random rotations were added to these images to produce two distinct 2D datasets for the study, each consisting of 5000 real images and lesion masks.

### Synthetic Images

Since the real 2D lesion images were extracted from a relatively small number of distinct subjects, the amount of non-redundant information in real datasets may be relatively limited. Thus, we employed two additional sets of 2D and 3D synthetic lesion images that were generated procedurally in-silico. In addition to being fully independent, synthetic images offer the advantage of having a more uniform background (i.e., unlike with real images, no other objects besides the lesion were present in the synthetic images).

We will describe our image synthesis methodology using the 2D case for brevity; in the 3D case, all aspects of methodology were symmetrically extended into the third dimension. First, a binary region representing the mask of the lesion was generated. To create a variety of mask shapes and sizes, a stochastic region growth algorithm was used, starting from 1 to 3 seeds that were randomly placed within a binary 48×48-pixel image; using a random number of initial seeds increased the variance of shape features, as confirmed in a post-hoc analysis. A random number (300-550) of region growth iterations was applied (Fig. 1a), and the resulting image was morphologically closed to remove small holes inside the mask.

**FIGURE 1.**
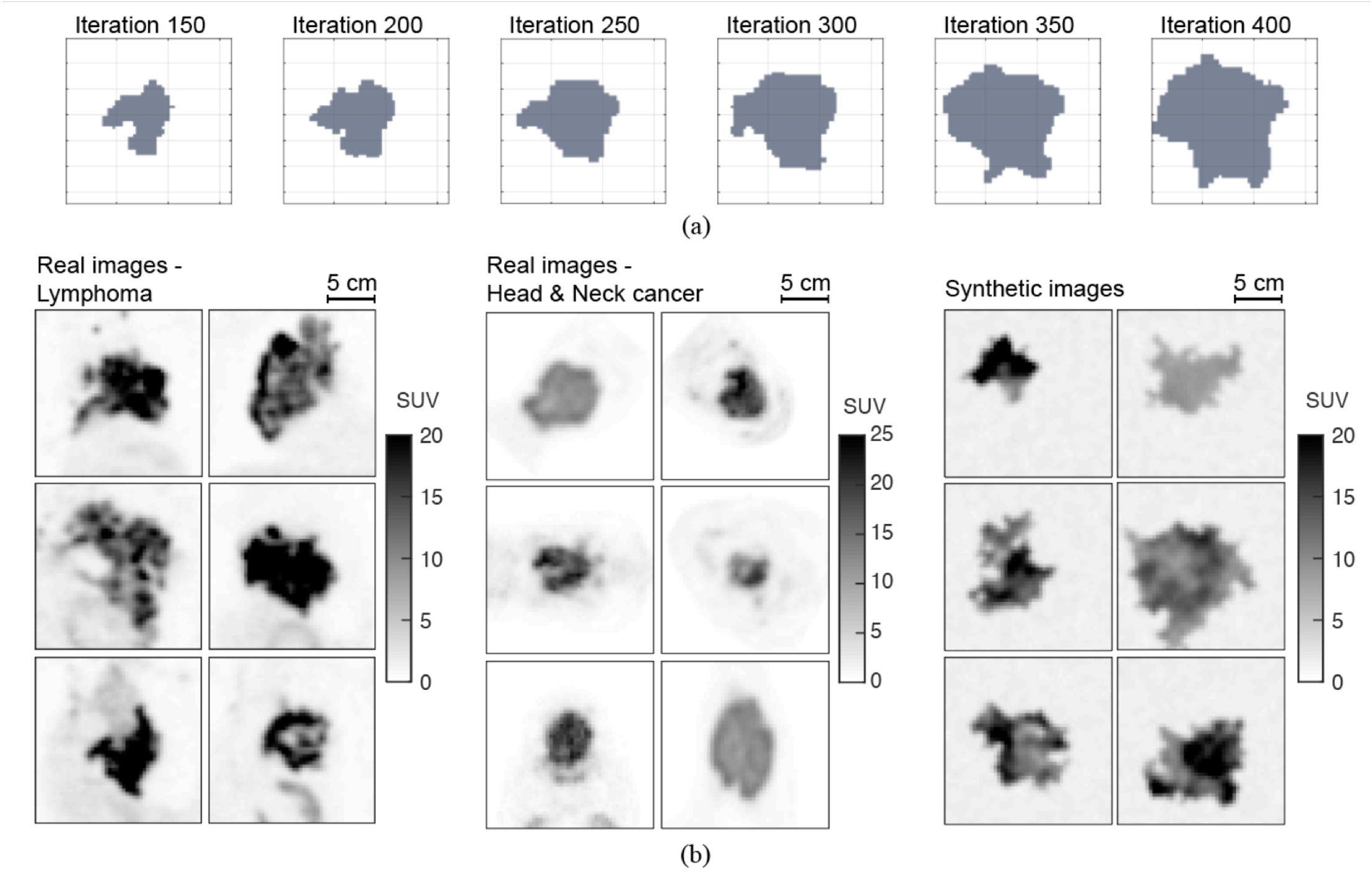
(A) Illustration of the region growth process utilized to generate random lesion shapes. (B) Examples of (left) acquired images of lymphoma and (middle) head and neck cancer tumors in comparison to (right) synthetic lesion images with different lesion intensities, shapes, and textures. The resolution and noise in synthetic images were matched to those of lymphoma images. The plotted synthetic and real images have the same dimensions (48×48 pixels) and isotropic pixel size (3.64 mm)^2^.

The lesion texture was created by generating a random Perlin pattern *(26)* and masking it using the generated mask. The pixel intensities were set to represent PET SUVs, and were scaled to vary between SUV_min_, chosen randomly between 2 and 7 for each image, and SUV_max_, chosen randomly between 9 and 30; SUV values in the background were set to 1.5. Magnitude-independent Gaussian noise (sigma = 0.15 SUV units) was added everywhere in the image, and spatial Gaussian smoothing (sigma = 0.85 pixels or 3.1 mm) was applied to the entire simulate resolution blurring. The isotropic pixel size (3.64 mm)^2^, resolution, and noise values were set to match those of the clinical images acquired at our center.

The SUV values and lesion sizes in the synthetic images were set to be similar to those in the real lymphoma images (Fig. 1b). A total of 5000 synthetic images were generated per each of 2D and 3D datasets.

### Radiomic Features

A set of 16 intensity, shape, and texture features, as defined by the Image Biomarker Standardization Initiative (IBSI) *(27)*, was selected for this study. The features and their statistics computed from the real images of lymphoma and synthetic 2D images are given in Table I. In both datasets, all 5000 images were used to compute the statistics. The choice of features was based on their simplicity, interpretability and frequency of use in research and clinical practice.

**TABLE 1.** Radiomic features computed from the real and synthetic 2D images and their statistics.

| Feature name | Real images (Lymphoma) |  | Synthetic images |  |
| --- | --- | --- | --- | --- |
|  | Median<br>(Q1, Q3) | (min, max) | Median<br>(Q1, Q3) | (min, max) |
| *Intensity features* |  |  |  |  |
| Maximum | 15.689<br>(11.081, 20.334) | (1.887, 40.003) | 16.948<br>(12.509, 21.276) | (6.839, 28.301) |
| Mean | 8.398<br>(5.705, 10.759) | (1.121, 24.158) | 11.026<br>(8.563, 13.405) | (4.859, 18.918) |
| Variance | 8.252<br>(3.534, 16.048) | (0.026, 78.698) | 5.645<br>(2.683, 9.871) | (0.597, 29.730) |
| COV | 0.337<br>(0.285, 0.402) | (0.067, 0.915) | 0.212<br>(0.184, 0.242) | (0.107, 0.425) |
| *Shape features - size* |  |  |  |  |
| Area | 2769.166<br>(1391.208, 4968.600) | (119.246, 14680.557) | 6121.315<br>(4213.373, 8347.248) | (808.226, 13236.351) |
| Convex area | 3497.835<br>(1647.192, 6181.809) | (119.246, 17474.762) | 7282.917<br>(5031.320, 9954.918) | (833.931, 16724.832) |
| Max diameter | 88.864<br>(61.018, 116.537) | (13.124, 199.005) | 116.537<br>(97.942, 136.294) | (38.003, 195.343) |
| Perimeter | 207.373<br>(123.394, 307.312) | (29.120, 972.379) | 381.879<br>(308.195, 457.694) | (99.422, 690.074) |
| *Shape features - irregularity* |  |  |  |  |
| Sphericity | 0.797<br>(0.704, 0.861) | (0.341, 0.968) | 0.694<br>(0.653, 0.734) | (0.448, 0.899) |
| Elongation | 0.604<br>(0.479, 0.731) | (0.126, 1.000) | 0.746<br>(0.657, 0.826) | (0.322, 0.997) |
| Solidity | 0.892<br>(0.776, 0.941) | (0.187, 1.000) | 0.846<br>(0.818, 0.870) | (0.617, 0.969) |
| Extent | 0.613<br>(0.509, 0.684) | (0.102, 0.846) | 0.589<br>(0.551, 0.626) | (0.393, 0.789) |
| *Texture features* |  |  |  |  |
| Contrast | 17.045<br>(12.408, 23.094) | (0.916, 67.663) | 8.586<br>(7.327, 10.148) | (4.025, 23.464) |
| Energy | 0.006<br>(0.005, 0.008) | (0.003, 0.528) | 0.008<br>(0.007, 0.011) | (0.004, 0.044) |
| Homogeneity | 0.362<br>(0.327, 0.404) | (0.228, 0.996) | 0.439<br>(0.413, 0.465) | (0.297, 0.605) |
| Entropy | 7.803<br>(7.433, 8.070) | (2.216, 8.791) | 7.371<br>(7.112, 7.590) | (5.518, 8.225) |

Intensity features are in SUV units, Area and Convex area are in mm<sup>2</sup>, Max diameter and Perimeter are in mm. Texture features were computed from the gray-level co-occurrence matrix (GLCM).

As per IBSI, the 4 intensity features describe first order pixel value statistics. The coefficient of variation (COV), often used as a measure of lesion heterogeneity *(3, 28)*, was computed as the ratio of the standard deviation to the mean.

The shape features include 4 descriptors of size and 4 descriptors of the shape irregularity. These features do not take into account the pixel intensities or their spatial distributions. Convex area was defined as the area of the convex envelope of the mask. Solidity is the ratio of lesion’s area to the convex area. Extent was defined as the ratio of lesion’s area to that of the axis-aligned bounding rectangle.

Texture features are represented by 4 second-order Haralick features computed from the gray level co-occurrence matrix (GLCM, 2.5D, merged); the pixel intensities were quantized using the constant bin number technique (32 bins). The ISBI-recommended constant bin size method was not used to minimize the interaction between the lesion intensity and texture. The voxel dimensions were specified to be isotropic for GLCM computation.

Table I demonstrates that the values features extracted from the synthetic images all fall within the min– max ranges of corresponding features computed from real lymphoma images. All features except perimeter and sphericity were computed using the IBSI-compliant SERA radiomics software *(29, 30)*. All features were extracted within lesion masks without image re-sampling. The masks were pre-processed to contain a single connected region without holes. The intensity features were computed using the original SUV units, and texture features used uniformly-discretized voxel intensities, with minimum and maximum intensities mapped to 1 and 32, respectively.

For the 2D and 3D image sets, all features were calculated in 2D and 3D, respectively (i.e. using the native image dimensionality).

### Tested Neural Net Architectures

We trained and tested several standard convolution-nonlinearity-pooling (CNP) architectures with an increasing number of convolutional layers. The hyper-parameters of the networks are specified in Table II, where the CN-2D-*X* abbreviations denote different CNP networks, and *X* is the number of convolutional layers; each convolutional layer consisted of 8 filters. The “Num. features” column contains the number of flattened features entering the final dense layer. The rectified linear unit (ReLU) nonlinearity was used throughout each CNP network, and max-pooling was used as the downsampling operation. After the flattening layer, all networks included one dense layer with 16 nodes, followed by a regression output layer. All parameters were trained. Since the number of max-pooling layers, flattened features, and dense layer nodes were fixed, the number of trainable parameters depended solely on the number of convolutional layers. In all convolutional filters, isotropic kernels of the size 5×5 were used; the kernel size of max-pooling layers was 2×2. The 3D CNP networks had the same structure as the 2D CNP networks.

**TABLE 2.**
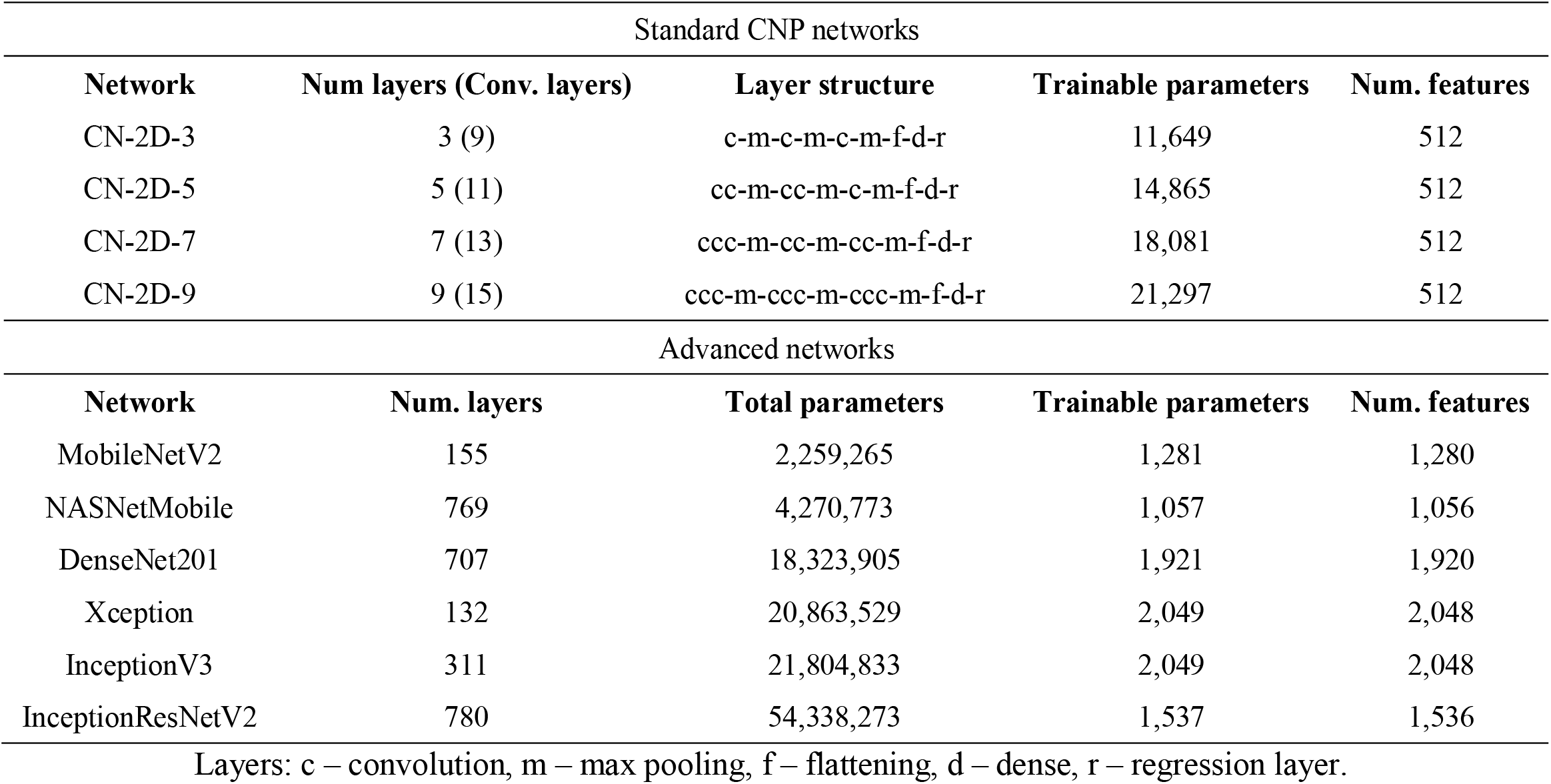
Parameters of the standard CNP and advanced ImageNet CNNs.

| Standard CNP networks |  |  |  |  |
| --- | --- | --- | --- | --- |
| Network | Num layers (Conv. layers) | Layer structure | Trainable parameters | Num. features |
| CN-2D-3 | 3 (9) | c-m-c-m-c-m-f-d-r | 11,649 | 512 |
| CN-2D-5 | 5 (11) | cc-m-cc-m-c-m-f-d-r | 14,865 | 512 |
| CN-2D-7 | 7 (13) | ccc-m-cc-m-cc-m-f-d-r | 18,081 | 512 |
| CN-2D-9 | 9 (15) | ccc-m-ccc-m-ccc-m-f-d-r | 21,297 | 512 |
| Advanced networks |  |  |  |  |
| Network | Num. layers | Total parameters | Trainable parameters | Num. features |
| MobileNetV2 | 155 | 2,259,265 | 1,281 | 1,280 |
| NASNetMobile | 769 | 4,270,773 | 1,057 | 1,056 |
| DenseNet201 | 707 | 18,323,905 | 1,921 | 1,920 |
| Xception | 132 | 20,863,529 | 2,049 | 2,048 |
| InceptionV3 | 311 | 21,804,833 | 2,049 | 2,048 |
| InceptionResNetV2 | 780 | 54,338,273 | 1,537 | 1,536 |

Layers: c – convolution, m – max pooling, f – flattening, d – dense, r – regression layer.

Additionally, several advanced ImageNet-pretrained CNN architectures were tested that included non-standard computation blocks and connections (Table II). The network graphs, weights, and biases were downloaded from the TensorFlow pre-trained model repository. The total number of parameters ranged from ∼2M to ∼54M. The pre-trained head (dense layers) of each network was removed, and a single new regression layer was added and trained, with the rest of the network frozen. The resulting number of trainable parameters was between 1057 and 2049, depending on the number of flattened features after the last convolutional layer. The networks were implemented in Python using the Keras module within TensorFlow v.2.2.

### Neural Net Training

Inputs to CNP and advanced networks were the real or synthetic SUV images, and target variables were the normalized values of radiomic features. The features were normalized by subtracting the mean and dividing by one standard deviation. One feature was tested at a time, i.e. each network only had one regression output. The networks were trained in end-to-end using the AdaGrad algorithm, with the base learning rate set to 0.01, decay rate set to 0, and initial accumulator value set to 0.1. The minimized loss function was the mean absolute error between predicted and ground truth values of radiomic features. Training was performed for 200 epochs, in mini-batches of 32 images. Out of 5000 in each dataset, 4000 were used for training, 500 for validation, and 500 for testing.

### Test Procedures and Metrics

Test sets of 500 images were used to assess the efficacy of CNP and advanced networks in learning radiomic features. Two metrics were used to quantify prediction error: 1) the normalized mean absolute error (nMAE):

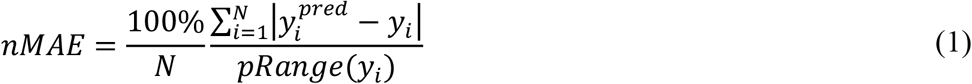

where *N* is the number of test samples, *y*_*i*_ is the true feature value, 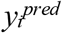 is the predicted feature value, and pRange is the percentile range (2.5–97.5); 2) Spearman’s correlation coefficient *ρ* between the ground truth and network-predicted feature values. The values of nMAE and *ρ* were computed on the test sets, for all tested datasets, features, and networks, as reported below.

To analyze the sample complexity for different features, we trained the CNP networks using different numbers of synthetic 2D image samples, ranging from 100 to 4500. The test loss and the difference between the training and test loss (i.e. train-test generalization) were measured as functions of the number of samples. Two additional tests were performed on the synthetic 2D dataset to aid the interpretation of results: 1) binary masks were tested as CNN inputs to investigate the effect of contrast on learning of shape features; 2) a dataset with fixed lesion size was tested to prevent networks from using size as a proxy for shape. We briefly report the results of these auxiliary tests.

## RESULTS

### Feature Prediction Errors From Real and Synthetic Images

Radiomic feature prediction errors on the lymphoma, head and neck, and synthetic test sets are plotted in Figures 2a, 2b, and 2c, respectively, for 2D CNP networks. The plots demonstrate a similar ranking of errors in real and synthetic images. In all datasets, the lowest values of nMAE were measured with size features (Area, Convex area, Max diameter, Perimeter), and with the Mean and Maximum intensity. This shows that the lesion intensity and size were the easiest features to learn for the CNNs. On the other hand, the prediction errors were 3-4 times higher for the shape irregularity features — Sphericity, Solidity, Elongation, and Extent. Notably, while Area and Convex area were predicted with high accuracy, their ratio defined as Solidity was predicted poorly.

**FIGURE 2.**
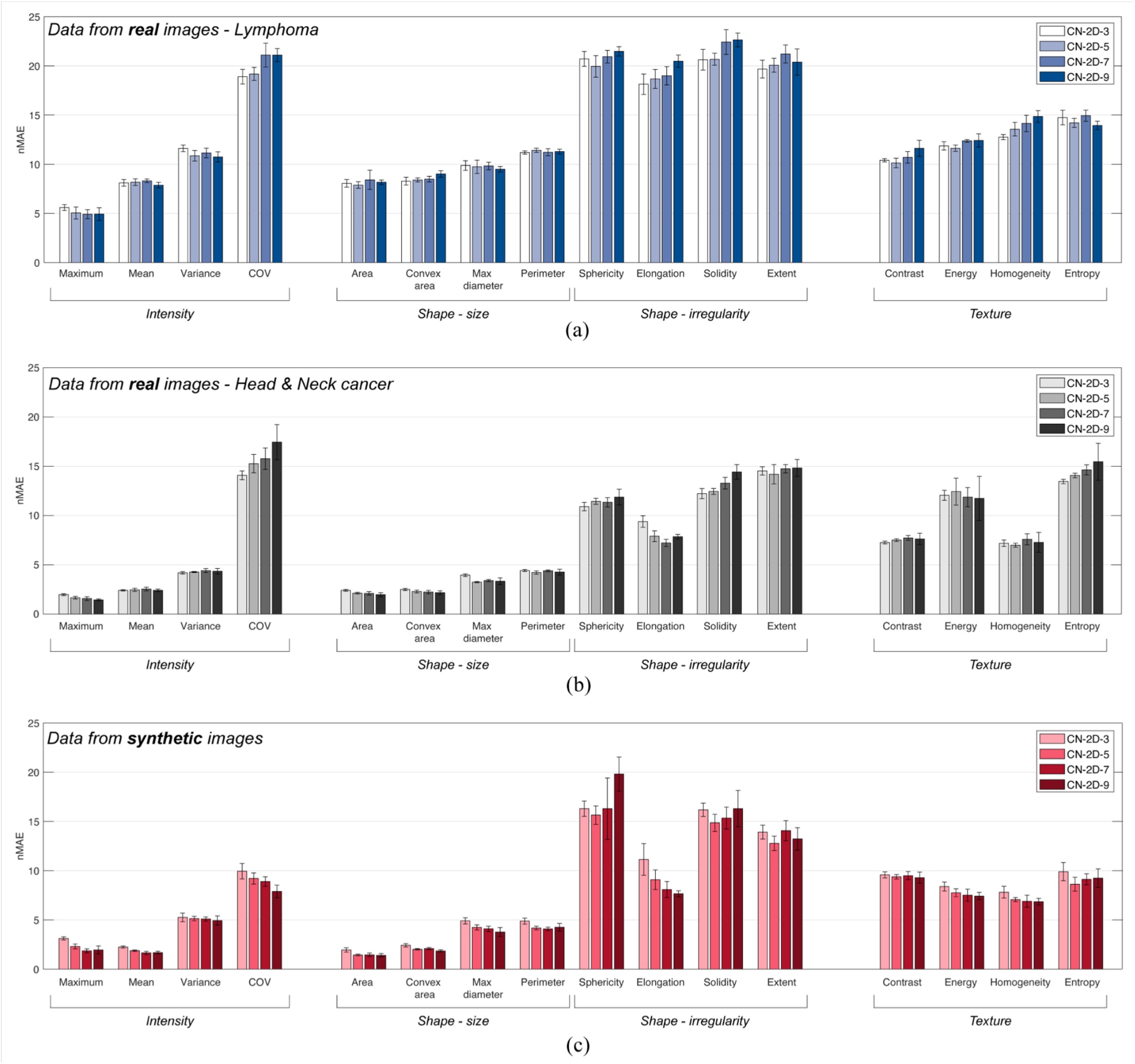
Radiomic feature prediction errors (nMAE) for the 2D CNP networks. (A) Data obtained with real lymphoma images. (B) Data obtained with real head and neck cancer images. (C) Data obtained with synthetic lesion images. The mean values and standard deviations were measured using 5 independent training trials.

Results obtained with 3D CNP networks and 3D synthetic images exhibit similar trends (available in Supplemental Figure 1 and Supplemental Table 1). The intensity and size features were predicted with relatively low errors, while shape features were predicted with high errors.

Representative scatter plots of true versus predicted feature values, obtained with real lymphoma images and the CN-2D-5 network, are plotted in Fig. 3. The scatter plots demonstrate that the high nMAE values for shape irregularity features did not originate from outliers or biases. Indeed, the data points for Sphericity and Solidity are substantially more scattered around the identity line compared to features like the Maximum intensity, Area, and Contrast. The same was observed in the corresponding scatter plots for synthetic images (not shown).

**FIGURE 3.**
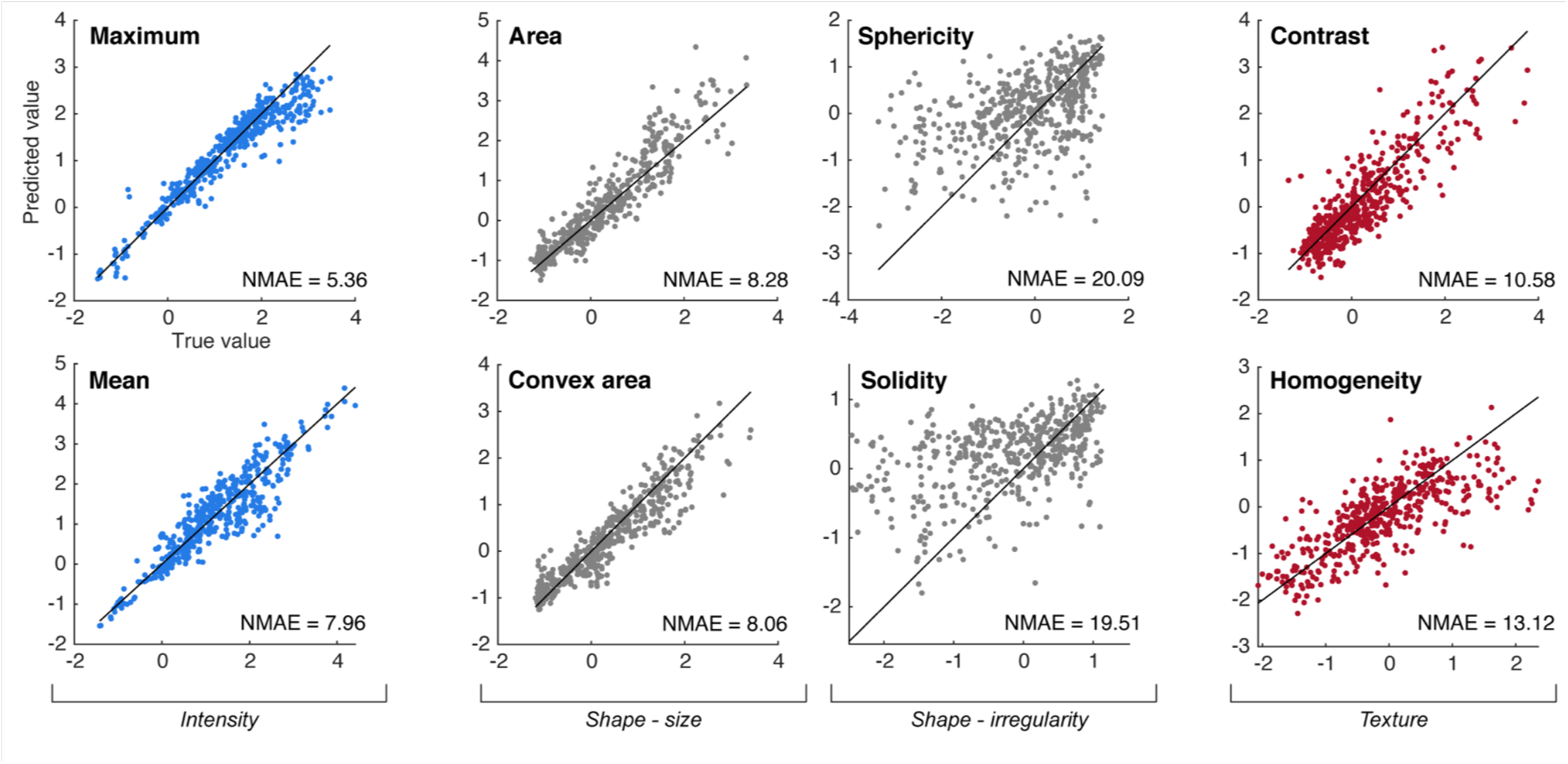
Predicted feature values from lymphoma images (normalized, y-axes) plotted against true feature values (normalized, x-axes) in the test set of 500 samples, for the CN-2D-5 network. The identity line is plotted in solid black color.

Generally, features predicted from real images had higher errors than those predicted from synthetic images. This likely reflects a more limited information content in the real 2D datasets (originating from a limited number of unique subjects). A related finding is that with real images the test errors generally increased with additional convolutional layers, implying the CNNs were overfitting the training data (Figs. 2a and 2b). In contrast, with synthetic images the prediction performance either improved, or remained the same with added convolutional layers (Fig. 2c).

Having confirmed that we observe the same trends in the real and synthetic datasets, with the latter yielding lower feature prediction errors, we only use the synthetic 2D images in the rest of the analysis.

### Spearman’s Correlation Coefficients

Spearman’s rank correlation coefficients *ρ* between predicted and true feature values for 2D CNP networks are listed in Table III; greater values correspond to better prediction performance. The given mean values and standard deviations were measured using 5 independent CNN training trials. Where omitted, the standard deviation was less than 0.01. The relative standing of features in terms of *ρ* was similar to that of nMAE: Sphericity, Solidity, and Extent had distinctly and significantly lower *ρ* values compared to other features. The findings were similar when an extended set of 36 features was examined (available in Supplemental Table 2).

**TABLE 3.** Spearman’s correlation coefficients *(ρ)* between predicted and true feature values (2D CNP networks).

| Feature name | CN-2D-3 | CN-2D-5 | CN-2D-7 | CN-2D-9 |
| --- | --- | --- | --- | --- |
| *Intensity features* |  |  |  |  |
| Maximum | 0.99 | 1.00 | 1.00 | 1.00 |
| Mean | 0.99 | 1.00 | 1.00 | 1.00 |
| Variance | 0.98 | 0.98 | 0.98 | 0.98 |
| COV | $0.86 \pm 0.03$ | $0.88 \pm 0.01$ | $0.89 \pm 0.01$ | $0.91 \pm 0.02$ |
| *Shape features - size* |  |  |  |  |
| Area | 1.00 | 1.00 | 1.00 | 1.00 |
| Convex area | 0.99 | 0.99 | 0.99 | 1.00 |
| Max diameter | 0.97 | 0.98 | 0.98 | 0.98 |
| Perimeter | 0.97 | 0.98 | 0.98 | 0.98 |
| *Shape features - irregularity* |  |  |  |  |
| Sphericity | **$0.65 \pm 0.03$** | **$0.68 \pm 0.04$** | **$0.66 \pm 0.13$** | **$0.54 \pm 0.07$** |
| Elongation | $0.82 \pm 0.05$ | $0.89 \pm 0.03$ | $0.91 \pm 0.02$ | $0.92 \pm 0.01$ |
| Solidity | **$0.58 \pm 0.04$** | **$0.67 \pm 0.04$** | **$0.65 \pm 0.04$** | **$0.62 \pm 0.09$** |
| Extent | **$0.69 \pm 0.04$** | **$0.75 \pm 0.03$** | **$0.69 \pm 0.04$** | **$0.73 \pm 0.04$** |
| *Texture features* |  |  |  |  |
| Contrast | $0.87 \pm 0.01$ | $0.87 \pm 0.01$ | $0.87 \pm 0.01$ | $0.87 \pm 0.02$ |
| Energy | $0.83 \pm 0.02$ | $0.85 \pm 0.01$ | $0.86 \pm 0.02$ | $0.86 \pm 0.01$ |
| Homogeneity | $0.90 \pm 0.01$ | $0.91 \pm 0.01$ | $0.92 \pm 0.01$ | $0.92 \pm 0.01$ |
| Entropy | $0.84 \pm 0.03$ | $0.88 \pm 0.02$ | $0.85 \pm 0.01$ | $0.86 \pm 0.03$ |

The three lowest (worst) values in each column are highlighted in bold.

### Train and Test Loss Analysis

To further investigate the high prediction errors for the shape irregularity features, training and test losses were inspected as functions of the training epoch. For illustration, training and test losses for the CN-2D-5 network trained on synthetic data are plotted in Fig. 4. After 200 epochs, the test loss had converged for all tested features. The fastest convergence (i.e. achieved in a fewest number of epochs) was observed with Maximum/Mean intensity, Variance, and size-related features such as Area and Volume. Features quantifying shape irregularity and COV had the slowest convergence.

**FIGURE 4.**
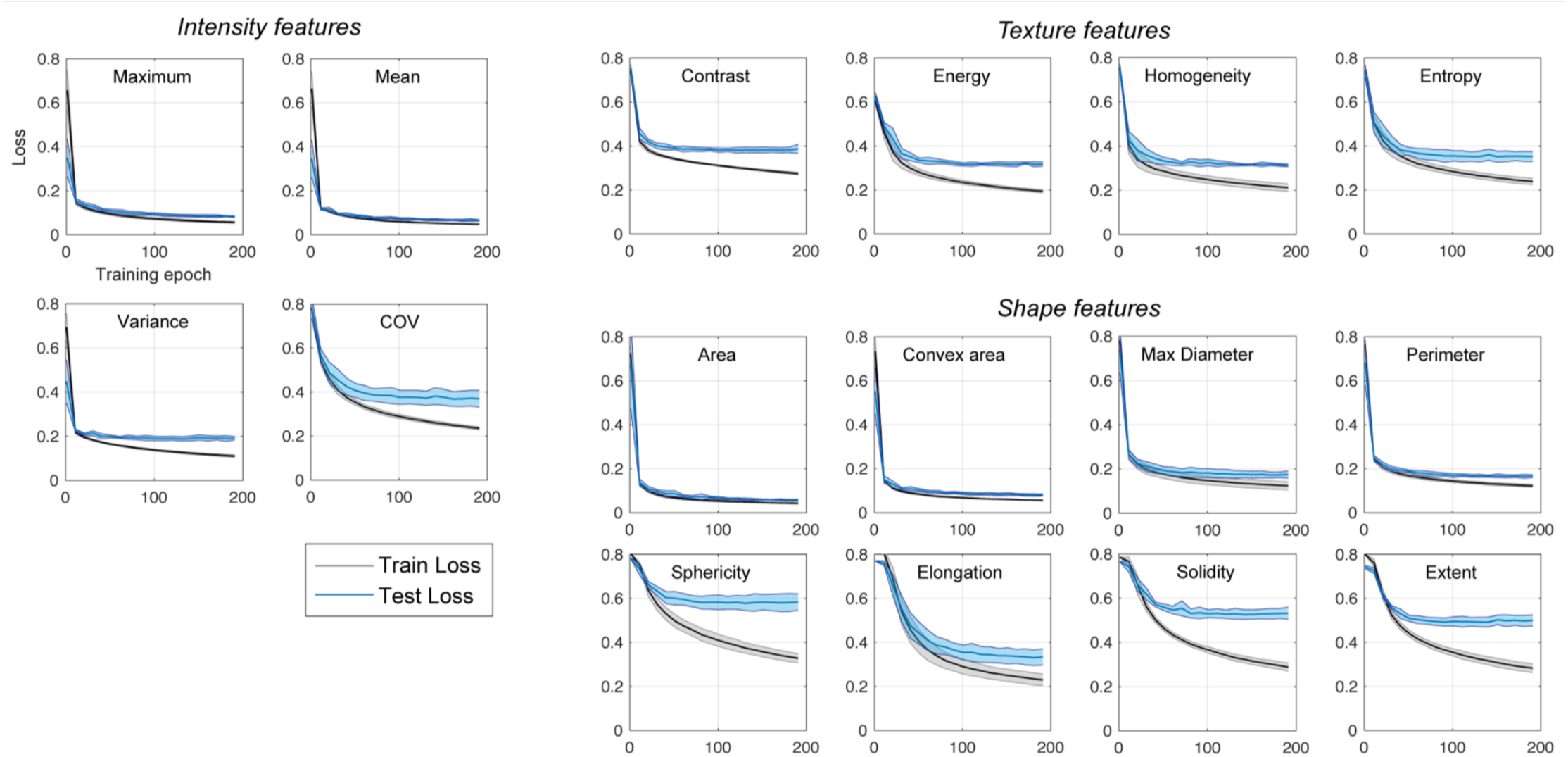
Training and test losses plotted against the training epoch for the CN-2D-5 network. The mean and standard deviation of loss values from 5 independent training trials are plotted.

With the shape irregularity features (Sphericity, Solidity end Extent), there was also a marked difference between the training and test losses. The relatively high training loss for these features indicates that the networks were less effective at approximating the respective functions. On the other hand, the even higher test loss indicates that the networks did not generalize well from the training to test sets.

### Sample Complexity Analysis

The generalization capacity of a CNN can be assessed from the difference between the training and test loss; sample complexity represents the number of training samples required to achieve good generalization in a broad sense. We measured the train-test loss difference with the CN-2D-3 network (simplest network tested) for a representative group of features, using various numbers of synthetic training images (Fig. 5).

**FIGURE 5.**
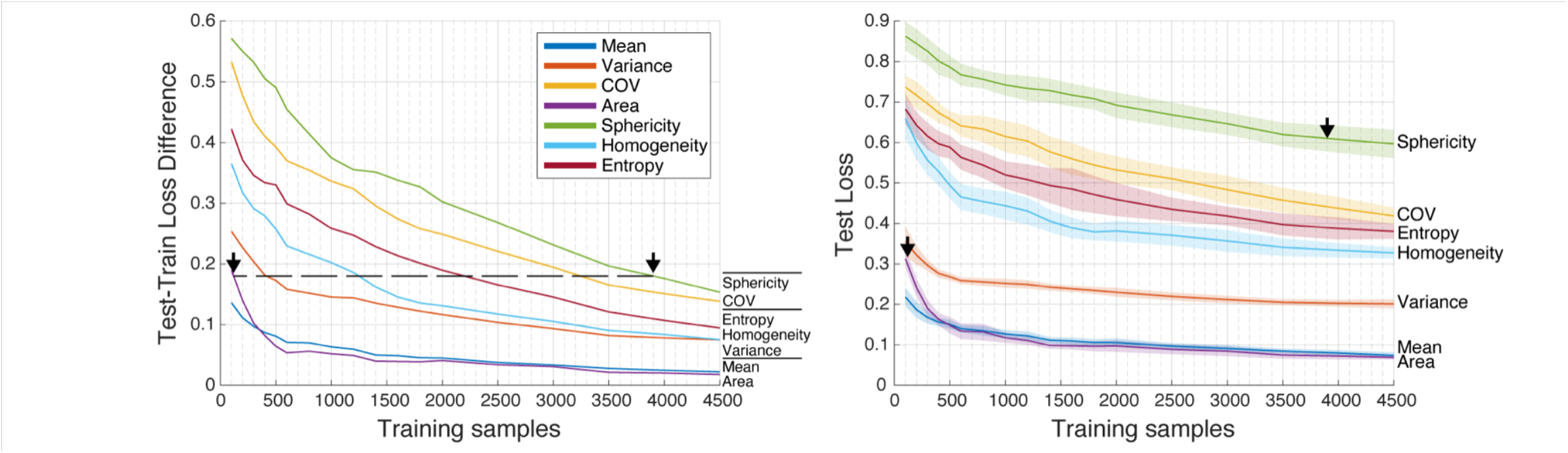
Left: difference between the train and test loss plotted against the number of training samples; arrows indicate the same value of the difference for Area and Sphericity. Right: test loss plotted against the number of training samples. Arrows indicate the values of test loss for Area and Sphericity achieved at the same level of generalization. The mean and standard deviations of loss values from 10 independent trials are plotted.

The graphs demonstrate the significantly different sample complexities for different features. In the extreme case, to achieve the same level of generalization, Area required ∼100 samples, and Sphericity required 3900 samples. Note that the corresponding test loss values for Area and Sphericity were 0.3 and 0.6, respectively, i.e. a similar generalization capacity between two features does not imply a similar prediction error.

### Additional Tests with CNP Networks

Using binary masks as inputs to the CNP networks (instead of SUV images) to predict the shape features resulted in the reduction of nMAE by approximately 20% for the Sphericity, Solidity and Extent features. The relative prediction errors for these features remained to be the highest.

When the lesion size was set to be constant in the synthetic images (though new dataset generation), the errors of shape feature predictions were similar to those plotted in Fig. 2. This may indicate that, when predicting shape features, our networks did not utilize any possible correlations between the lesion shape and size.

Increasing the convolutional kernel sizes in the CNP networks resulted in a worse performance by approximately 20%, compared to the results shown in Figure 2. We varied the AdaGrad step size between 1 and 0.0001, and the results were found to be similar. Likewise, using a smaller (16) or larger (64) batch size did not change the results significantly.

### Advanced Networks

Radiomic feature prediction errors obtained with the synthetic 2D images and advanced networks are plotted in Fig. 6. The intensity and size features were predicted with higher errors compared to the CNP networks trained from scratch. Advanced networks with a greater number of trainable parameters or layers did not produce lower prediction errors. On the contrary, the intensity features were predicted best with the MobileNetV2 network, which had the fewest number of parameters. Inspection of the true-vs-predicted value scatter plots (not shown) confirmed that the high prediction errors were distributed uniformly among the test samples and not originate from a few outliers.

**FIGURE 6.**
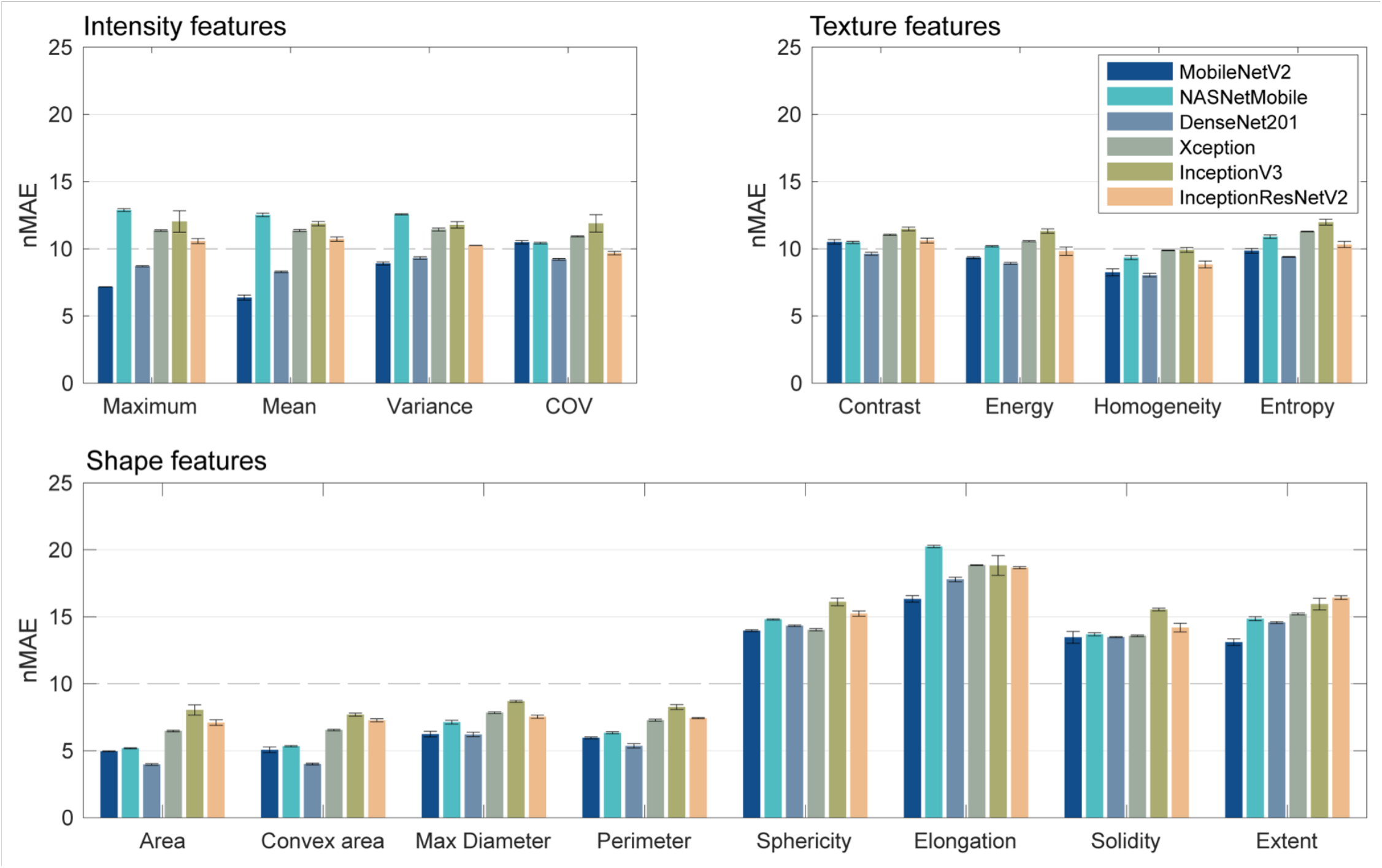
Feature prediction errors (nMAE) obtained with the advanced networks on synthetic lesion images. The mean values and standard deviations were measured using 3 independent trials of the regression layer training.

The standard deviations of errors with the advanced networks were markedly lower compared to those of the CNP networks, due to the frozen parameters being constant between different trials. Likewise, the training and test loss of advanced networks converged on average within the first 20 epochs, much faster compared to the CNP networks. We found that the test loss closely followed the training loss with most features, although the difference between the training and test losses was again greater with the shape irregularity features.

Spearman’s rank correlation coefficients *ρ* between predicted and true feature values for the advanced networks are given in Table IV. The given mean values and standard deviations were measured using 3 independent trials of the regression layer training. Where omitted, the standard deviation was less than 0.01. The ranking of features and networks in terms of *ρ* was similar to that obtained with nMAE.

**TABLE 4.**
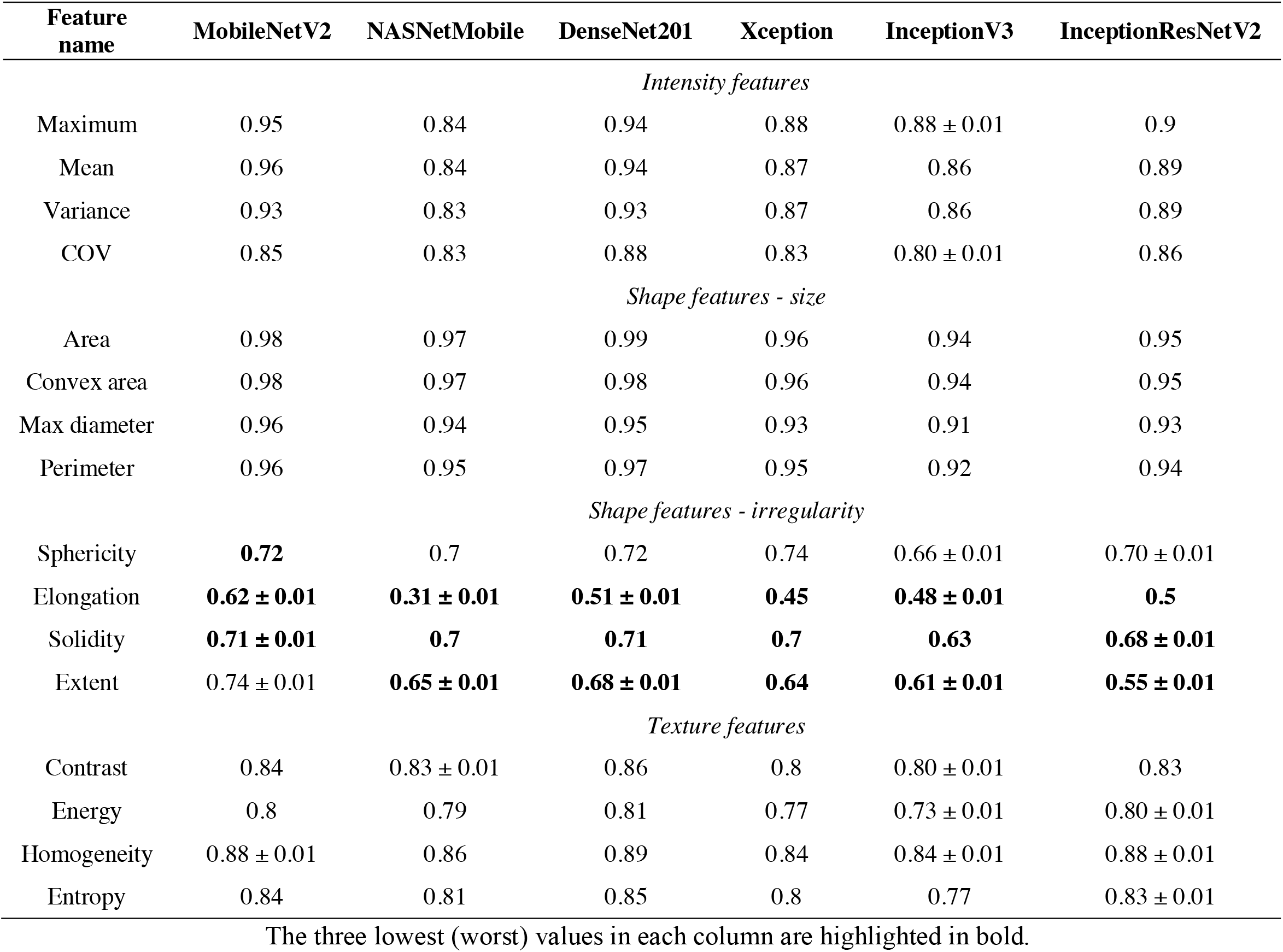
Spearman’s correlation coefficients *(ρ)* between predicted and true feature values (Advanced networks).

## DISCUSSION

We have directly quantified the relative expressive power of standard CNN architectures with respect to standardized intensity, shape, and texture features commonly used in oncological imaging. In two real and two synthetic datasets, we found that features quantifying lesion size as well as maximum and mean intensities exhibited lowest prediction errors. On the other hand, features quantifying shape irregularity had highest prediction errors, and generalized poorly from the training to test sets. Given that tumor shape has been found to be a significant predictor of clinical outcomes *(5, 31)*, this finding may bear significant implications for the use of CNNs in clinical prediction tasks. For example, CNNs that are trained to predict progression-free survival from tumor images, may preferentially learn to leverage the intensity and size information, while the shape-irregularity information may be under-utilized.

In addition to standard CNNs trained end-to-end, we tested several ImageNet pre-trained advanced networks that were fine-tuned on our data. We found that all radiomic features predicted by advanced networks had high errors, higher than those obtained with standard CNNs, implying that radiomics-related information is poorly represented in the high-level feature output layers of ImageNet pre-trained networks. The errors were highest for the shape features, mirroring findings with the standard CNNs. Based on these observations, we conclude that simple CNNs trained end-to-end on domain-specific images should capture radiomic features better than advanced networks pre-trained on large image sets like ImageNet. This likely happens because the latter layers in advanced CNNs become over-specialized when trained to classify ImageNet images: the best performance among the advanced networks was obtained with MobileNetV2, which was the simplest network in terms of the number of layers and parameters.

Sample complexity analysis showed that intensity and size metrics required around 100-500 training samples to achieve good train-test generalization. On the other hand, shape irregularity features required around 2000-4000 training samples. Hence, a relatively large number of examples is required for CNNs to capture the shape-related information from the images. In contrast, medical imaging studies that use CNNs often have far fewer than 1000 training samples — typically the number of samples is on the order of 100 or less, particularly for PET studies (according to the Cancer Imaging Archive, https://www.cancerimagingarchive.net) *(32)*. In studies with tens or hundreds of samples, CNNs may only be able to implicitly learn “easier” features related to the intensity and size (image augmentation may help to alleviate this issue).

We hypothesize that high prediction errors for some features may be attributed to two factors. First, the tested networks lacked direct ability to capture global context, which may be important for capturing global shape properties. Designing and using CNNs that can capture the global context and have larger perceptive fields may lead to a better implicit learning of shape properties. Second, the high prediction errors could have originated from the limited ability of CNNs to approximate ratio-type features or functions (such as COV, sphericity, solidity and extent). For example, solidity is a ratio of area and convex area, both of which were predicted with a much lower error compared to solidity. Including a non-standard division operation in the network graph, or adding the reciprocal image as an input, may improve prediction performance. Alternatively, features with high prediction errors can be added explicitly as auxiliary variables to the dense layers in the “heads” of the networks, or as additional input channels. We propose that making these modifications to existing and previously published models for image-based diagnosis may improve the performance of the models. An interesting direction of future research is to compare the performance of standard and radiomics-augmented neural networks in tasks that predict clinical metrics or outcomes.

Among other findings, there was an unexpectedly small improvement in the prediction error with added network depth. It is of interest to explore how the width of the network, i.e. the number of filters or channels in the convolutional layers, affects prediction errors: shallower and wider CNNs may perform as well or better than deeper networks in medical imaging applications. Recent theoretical studies suggest that the expressive power of neural networks grows faster with added depth than with added width *(33, 34)*. However, this may or may not apply to functions that represent low-level image features.

A limitation of our study is that the tests were performed only on PET images, real and synthetic. However, we believe that our findings, particularly with shape features, should generalize to other modalities (since shape analysis does not utilize pixel intensities). It is of particular interest to reproduce our experiments on CT and MRI images, where larger datasets are available.

## CONCLUSIONS

Our work shows that conventional CNNs architectures readily learn first-order intensity and size-related radiomic features from less than 500 samples. On the other hand, features describing tumor heterogeneity (e.g. COV) and shape irregularity are difficult to learn, and require an order of magnitude more samples; the capacity of CNNs to learn texture features is intermediate. Therefore, CNNs may not be as effective as explicit radiomic features at capturing certain tumor properties. This is in fact more strongly the case for CNNs pretrained on image sets like ImageNet. In our view, the use of explicit radiomics and traditional machine learning techniques may not be properly discarded in favor of existing CNNs when it comes to medical image analysis, as the strengths of these two approaches appear to be complementary: a combination of the two approaches or appropriate next-generation deep networks are likely to produce improved results.

## Data Availability

The data used in this study contained protected patient health information and are only available under the respective IRBs. Parts of data not covered by IRBs can be made available upon request.

## ACKNOWLEDGMENT

This work was supported by the National Institutes of Health (NIH) / Canadian Institutes of Health Research (CIHR) Quantitative Imaging Network (QIN) Grant number 137993, CIHR Project Grant PJT-162216, and in part through computational resources and services provided by Microsoft and the Vice President Research and Innovation at the University of British Columbia. The authors also acknowledge Dr. Kerry Savage, Dr. Carlos Uribe, and Dr. Fereshteh Yousefirizi for helpful discussions and sharing of PET/CT images for this study.

