## Supplemental material for "Testing the Ability of Convolutional Neural Networks to Learn Radiomic Features"

**SUPPLEMENTAL DATA**

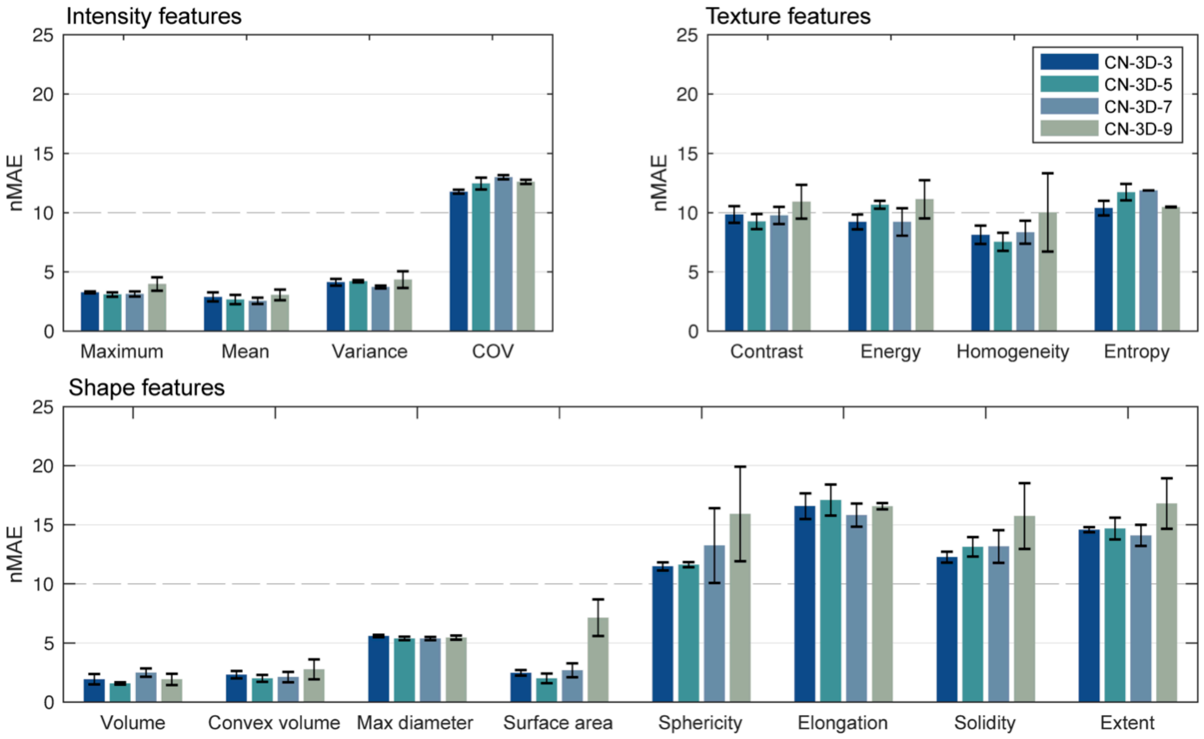


SUPPLEMENTAL FIGURE 1: Radiomic feature prediction errors (nMAE) for 3D CNP networks trained on synthetic 3D lesion images. The mean values and standard deviations were measured using 3 independent training trials. The following 2D features were replaced by their 3D equivalents: Area replaced with Volume, Convex area replaced with Convex volume, Perimeter replaced with Surface area.

SUPPLEMENTAL TABLE 1: Spearman's rank correlation coefficients (ρ) between predicted and true feature values for 3D CNP networks.

| **Feature name** | **CN-3D-3** | **CN-3D-5** | **CN-3D-7** | **CN-3D-9** |
|---|---|---|---|---|
| | | *Intensity features* | | |
| Maximum | 0.99 | 0.99 | 0.99 | 0.99 |
| Mean | 0.99 | 0.99 | 0.99 | 0.93 ± 0.11 |
| Variance | 0.99 | 0.99 | 0.99 | 0.99 |
| COV | 0.82 ± 0.01 | 0.79 ± 0.02 | 0.78 ± 0.01 | 0.79 ± 0.01 |
| | | *Shape features* | | |
| Volume | 1.00 | 1.00 | 0.99 | 1.00 |
| Convex volume | 0.99 | 0.99 | 0.99 | 0.99 ± 0.01 |
| Max diameter | 0.96 | 0.96 | 0.96 | 0.96 |
| Surface area | 0.99 | 0.99 | 0.99 | 0.91 ± 0.13 |
| Sphericity | 0.81 ± 0.01 | 0.80 ± 0.02 | 0.81 ± 0.03 | 0.81 ± 0.03 |
| Elongation | **0.61 ± 0.02** | **0.57 ± 0.07** | **0.66 ± 0.03** | **0.61 ± 0.02** |
| Solidity | **0.71 ± 0.03** | **0.70 ± 0.03** | **0.70 ± 0.06** | **0.66 ± 0.06** |
| Extent | **0.68 ± 0.01** | **0.67 ± 0.04** | **0.70 ± 0.05** | **0.62 ± 0.05** |
| | | *Texture features* | | |
| Contrast | 0.90 ± 0.01 | 0.91 ± 0.01 | 0.90 ± 0.02 | 0.90 ± 0.01 |
| Energy | 0.85 ± 0.02 | 0.82 ± 0.02 | 0.86 ± 0.03 | 0.80 ± 0.04 |
| Homogeneity | 0.92 ± 0.02 | 0.94 ± 0.01 | 0.93 ± 0.02 | 0.89 ± 0.07 |
| Entropy | 0.85 ± 0.01 | 0.82 ± 0.01 | 0.82 ± 0.02 | 0.85 ± 0.03 |

The three lowest (worst) values in each column are highlighted in bold. The mean values and standard deviations were measured using 3 independent CNN training trials. Where omitted, the standard deviation was less than 0.01. GLCM: gray level co-occurrence matrix.

SUPPLEMENTAL TABLE 2: Spearman's correlation coefficients (ϱ) between predicted and true feature values (2D CNP networks), for an extended set of features.

| **Feature name** | **CN-2D-3** | **CN-2D-5** | **CN-2D-7** | **CN-2D-9** |
|---|---|---|---|---|
| | | *Intensity features* | | |
| Maximum | 0.99 | 1.00 | 1.00 | 1.00 |
| Mean | 0.99 | 1.00 | 1.00 | 1.00 |
| Variance | 0.98 | 0.98 | 0.98 | 0.98 |
| COV | 0.86 ± 0.03 | 0.88 ± 0.01 | 0.89 ± 0.01 | 0.91 ± 0.02 |
| Skewness | 0.75 ± 0.04 | 0.83 ± 0.01 | 0.83 ± 0.05 | 0.86 ± 0.02 |
| Kurtosis | 0.49 ± 0.02 | 0.52 ± 0.03 | 0.45 ± 0.06 | 0.55 ± 0.08 |
| Median | 0.99 | 0.99 | 1.00 | 1.00 |
| 90th percentile | 1.00 | 1.00 | 1.00 | 1.00 |
| Mean absolute deviation | 0.97 | 0.98 | 0.97 ± 0.01 | 0.98 |
| | | *Shape features* | | |
| Area | 1.00 | 1.00 | 1.00 | 1.00 |
| Convex area | 0.99 | 0.99 | 0.99 | 1.00 |
| Max diameter | 0.97 | 0.98 | 0.98 | 0.98 |
| Perimeter | 0.97 | 0.98 | 0.98 | 0.98 |
| Sphericity | 0.65 ± 0.03 | 0.68 ± 0.04 | 0.66 ± 0.13 | 0.54 ± 0.07 |
| Elongation | 0.82 ± 0.05 | 0.89 ± 0.03 | 0.91 ± 0.02 | 0.92 ± 0.01 |
| Solidity | 0.58 ± 0.04 | 0.67 ± 0.04 | 0.65 ± 0.04 | 0.62 ± 0.09 |
| Extent | 0.69 ± 0.04 | 0.75 ± 0.03 | 0.69 ± 0.04 | 0.73 ± 0.04 |
| Centre of Mass Shift | 0.18 ± 0.04 | 0.19 ± 0.09 | 0.21 ± 0.08 | 0.25 ± 0.06 |
| Moran's I Index | 0.64 ± 0.04 | 0.63 ± 0.05 | 0.68 ± 0.07 | 0.72 ± 0.05 |
| Geary's C Measure | 0.60 ± 0.02 | 0.59 ± 0.02 | 0.58 ± 0.05 | 0.55 ± 0.05 |
| | | *Texture features* | | |
| GLCM Contrast | 0.87 ± 0.01 | 0.87 ± 0.01 | 0.87 ± 0.01 | 0.87 ± 0.02 |
| GLCM Energy | 0.83 ± 0.02 | 0.85 ± 0.01 | 0.86 ± 0.02 | 0.86 ± 0.01 |
| GLCM Homogeneity | 0.90 ± 0.01 | 0.91 ± 0.01 | 0.92 ± 0.01 | 0.92 ± 0.01 |
| GLCM Entropy | 0.84 ± 0.03 | 0.88 ± 0.02 | 0.85 ± 0.01 | 0.86 ± 0.03 |
| GLCM Dissimilarity | 0.90 | 0.90 ± 0.01 | 0.91 ± 0.01 | 0.90 ± 0.01 |
| GLCM Inverse Variance | 0.85 ± 0.01 | 0.86 ± 0.01 | 0.87 ± 0.02 | 0.88 ± 0.01 |
| GLCM Correlation | 0.59 ± 0.03 | 0.68 ± 0.08 | 0.59 ± 0.16 | 0.67 ± 0.14 |
| GLCM Cluster Tendency | 0.79 ± 0.03 | 0.82 ± 0.03 | 0.81 ± 0.05 | 0.78 ± 0.07 |
| GLCM Cluster Shade | 0.48 ± 0.05 | 0.56 ± 0.10 | 0.61 ± 0.02 | 0.59 ± 0.13 |
| GLCM Cluster Prominence | 0.76 ± 0.02 | 0.76 ± 0.03 | 0.74 ± 0.03 | 0.74 ± 0.03 |
| GLRLM Short Run Emphasis | 0.87 ± 0.01 | 0.88 ± 0.01 | 0.88 ± 0.02 | 0.87 ± 0.03 |
| GLRLM Long Run Emphasis | 0.88 ± 0.01 | 0.89 ± 0.01 | 0.89 ± 0.02 | 0.90 ± 0.01 |
| GLRLM Gray Level Nonuniformity | 0.75 ± 0.02 | 0.76 ± 0.01 | 0.79 ± 0.02 | 0.77 ± 0.10 |
| GLRLM Run Length Nonuniformity | 0.86 | 0.86 ± 0.03 | 0.87 ± 0.02 | 0.87 ± 0.04 |
| GLRLM Run Percentage | 0.99 | 1.00 | 1.00 | 1.00 |
| GLRLM Run Length Variance | 0.85 ± 0.01 | 0.87 ± 0.01 | 0.87 ± 0.01 | 0.87 ± 0.02 |

The mean values and standard deviations were measured using 5 independent CNN training trials. Where omitted, the standard deviation was less than 0.01. GLCM: gray level co-occurrence matrix; GLRLM: gray-level run-length matrix.
